# Motoric Cognitive Risk syndrome trajectories and incident dementia over 10 years

**DOI:** 10.1101/2023.06.22.23291741

**Authors:** Donncha S. Mullin, Danni Gadd, Tom C. Russ, Michelle Luciano, Graciela Muniz-Terrera

## Abstract

**Background:** Motoric Cognitive Risk (MCR) syndrome is a high-risk state for adverse health outcomes in older adults characterised by measured slow gait speed and self-reported cognitive complaints. The recent addition to the Lothian Birth Cohort 1936 of robust dementia outcomes enabled us to assess the prognostic value of MCR for dementia and explore the various trajectories of participants diagnosed with MCR.

**Methods:** We classified 680 community-dwelling participants free from dementia into non-MCR or MCR groups at mean [SD] age 76.3 [0.8] years. We used Cox and competing risk regression methods, adjusted for potential confounders, to evaluate the risk of developing all-cause incident dementia over 10 years of follow-up. Secondarily, we followed the trajectories for individuals with and without MCR at baseline and categorised them into subgroups based on whether MCR was still present at the next research wave, three years later.

**Results:** The presence of MCR increased the risk of incident dementia (adjusted HR 2.34, 95%CI 1.14-4.78, p=0.020), as did fewer years of education and higher depression symptoms. However, MCR has a heterogenous progression trajectory. The MCR progression subgroups each have different prognostic values for incident dementia.

**Conclusion:** MCR showed similar prognostic ability for dementia in a Scottish cohort as for other populations. MCR could identify a target group for early interventions of modifiable risk factors to prevent incident dementia. This study illustrates the heterogeneous nature of MCR progression. Exploring the underlying reasons will be important work in future work.

## Introduction

Dementia is a major global public health concern with no effective treatment. It is vital to focus on identifying the early predementia stage as this is when addressing modifiable risk factors and organising future care may be most effective at reducing the impact of dementia.^1^ Subjective cognitive complaints and slow walking speed are among the earliest reported findings in the pre-clinical stage of dementia, often detectable approximately 10 years before dementia diagnosis.^2^ Motoric cognitive risk (MCR) is a predementia syndrome defined as objective slow gait speed and subjective cognitive complaint in functionally independent individuals free of dementia.^3^ Diagnosing MCR is quick, inexpensive, and simple to do, which gives it great potential clinical utility. Diagnosing MCR could also assist research trials with cohort recruitment and ultimately contribute to a reduction in the prevalence of dementia. Given that approximately 50 million people worldwide live with dementia, a number projected to triple over the next 30 years,^4^ even a small reduction in incidence or delaying the age of onset could make a significant difference to patients, families and societies globally.^5,6^

First defined by Verghese in 2013^7^, MCR demonstrates good prognostic value as a high-risk state for developing dementia in cohorts worldwide, but this has not yet been studied in a Scottish cohort.^8–11^ A robust clinical dementia identification process using electronic medical record linkage was recently completed in the Scottish ageing cohort, the Lothian Birth Cohort 1936 (LBC1936).^12^ This process identified 118 out of 865 participants (13.6%) who were diagnosed with probable all-cause dementia, using the International Classification of Diseases-11 criteria.^13^ This recent addition to the LBC1936 makes it possible for the first time to assess the prognostic value of MCR for dementia in this Scottish cohort. However, MCR is not an inevitable prelude to future dementia. The first study examining the transient nature of MCR found that different clinical characteristics were associated with different MCR subtypes (e.g. stable, new, transient), but that MCR is associated with incident dementia regardless of subtype ^14^. Understanding the trajectories of those diagnosed with MCR is crucial to fully appreciate its clinical utility as a predictor of dementia.

Our study has the following aims:

1. to assess the prognostic value of MCR for incident dementia in a Scottish cohort of older adults;
2. to explore the various trajectories of participants diagnosed with MCR.

## Methods

### Study design, setting and sample size

This longitudinal prospective study used data from the Lothian Birth Cohort 1936 (LBC1936) study, which has been described in detail previously.^15–17^ In summary, the LBC1936 recruited 1091 participants aged 70 years living in the Lothian region of Scotland, most of whom had completed an intelligence test at age 11 years. Waves of testing have been conducted every three years since then. Data are available for five waves (mean ages 70, 73, 76, 79 and 82 years). A sixth wave has recently finished – but data are not yet available – and a seventh wave is planned. Each wave consists of interviews, cognitive tests, questionnaires, blood tests, and physical measures, including gait speed measurement. At wave 2, participants were first asked for written consent for medical data linkage, which enabled the identification of dementia regardless of whether participants returned to later waves of the LBC1936 or not. LBC1936 has an almost equal sex split, and all participants are white. To minimise loss to follow-up between waves, the LBC1936 researchers re-contact those unable to attend a wave due to a temporary illness and see them at a later, more appropriate time.^16^ The information necessary for deriving MCR was first collected at wave 3 in LBC1936 (mean age 76 years, *n* = 697), which determined our starting sample size.

### Eligibility criteria

We excluded participants receiving a dementia diagnosis within one year of their MCR categorisation. This reduces the risk of detecting pre-existing rather than incident dementia when performing time-to-event analysis. We excluded one participant who did not give consent for medical data linkage. We excluded participants missing data in any MCR criteria.

### Outcome variable: incident dementia

Clinicians recently diagnosed dementia in the LBC1936 cohort based on the International Classification of Diseases-11 criteria.^12^ This multi-step process involved i) a thorough clinician review of the electronic health records of every LBC1936 participant that consented to medical data linkage, ii) clinician assessment when there were concerns about a participant’s cognitive function, and iii) a diagnostic review board meeting of dementia experts. As the process for identifying dementia relies on linked medical data rather than LBC1936 testing, participants who dropped out of the study after wave 3 still have a dementia outcome. This markedly reduces the risk of attrition bias.

### MCR

Our primary risk factor of interest was MCR, defined as originally proposed by Verghese.^7^ Using data previously collected in the LBC1936, we identified participants who fulfilled the following MCR criteria:

1. Slow gait measured over 6 metres: ≥ 1 SD slower than sex and age-matched mean speed.
2. Self-reported cognitive complaint: answered “Yes” to the question “Do you currently have any problems with your memory?”
3. Functional independence: <= 1.5 SD above the mean on the Townsend Disability Scale overall score (higher score equals greater disability).^18^
4. No dementia: does not self-report or have a formal diagnosis of dementia and scores at least 24 on the Mini-Mental State Examination (MMSE).^19^

For our secondary analysis, we followed the participants from wave 3 (our baseline) to wave 4 (three years later) to define subtypes of MCR: New MCR (no MCR at baseline but MCR after three years), Transient Improved MCR (MCR at baseline but no MCR after three years due to an improvement – no longer a slow walker or no longer reported cognitive complaint), Transient Impaired MCR (MCR at baseline but no MCR after three years due to a deterioration – no longer functionally independent), and Stable MCR (MCR at baseline and after three years). This approach builds on a recent analysis of MCR subtypes.^14^ We split the Transient MCR group into ‘improved’ and ‘impaired’ as these are markedly different outcomes and it was important not to pool them. Finally, we defined a separate group of people who never developed MCR, Never MCR (no MCR at baseline and no MCR after three years).

### Covariates

Based primarily on previously reported risk factors for MCR and dementia,^5,11,20–24^ we selected the following risk factors in our analysis: age, sex, years of education, body mass index (BMI [kg/m2]), smoking status (current/ex/never), occupational social status (non-manual/manual), depression symptoms (Hospital Anxiety and Depression Scale), and sedentary lifestyle (self-reported physical activity level). The presence of self-reported stroke, hypertension, cardiovascular disease, diabetes, Parkinson’s disease, arthritis, leg pain, or neoplasia was used to calculate a summary multimorbidity index (scored 0 to 8).^2^ Self-reported physical activity levels were categorised into “Low”, “Medium”, and “High”, as detailed in Appendix 2.

### Statistical methods

In our primary analysis, we summarised the baseline characteristics of participants with and without MCR using descriptive statistics. We used ANOVA (continuous variables) and Pearson χ^2^ tests or Fisher’s as appropriate (categorical variables) to assess characteristics associated with and without MCR. We used Kaplan-Meier estimates of survival functions to illustrate differences in dementia-free survival between participants with and without MCR. A log-rank test compared the cumulative survival rates between those with and without MCR. To determine the effect of baseline MCR on incident dementia over a maximum of 10 years follow-up, we used Cox proportional hazards models to compute adjusted hazard ratios (HR) with 95% confidence intervals (CI). To reduce bias in estimates of the influence of predictors, we also used the Fine-Gray competing risk method to estimate the risk of dementia when death was a competing risk.^25,26^ For both time-to-event analysis methods, person-time variables were obtained by calculating the time between the wave 3 assessment date (i.e., when MCR was first derived, our study’s baseline) and the earliest of the following: i) dementia diagnosis date, ii) death, or iii) 18^th^ August 2022 (i.e., the end of the LBC1936 dementia ascertainment period)^27^ if the participant remained alive and dementia-free throughout the study follow-up. The proportionality assumption of the models were examined graphically and statistically and found to be adequately met. All analyses are adjusted for age, sex, and education. Subsequent models adjusted for additional covariates. To account for the possibility that the findings may have been biased from missing data, we compared missing data distribution among participants with and without dementia. There is equal distribution. We also include a missing values map to illustrate the lack of any non-random missingness in the covariates (Appendix 1).

For our secondary analysis, we used the same statistical approaches as for our primary analysis when describing and comparing the characteristics of the MCR subgroups, and when doing time-to-event analysis. We also used Kaplan-Meier estimates of survival functions to illustrate differences in dementia-free survival between the MCR subgroups.

All analyses were performed in R version 4.0.2, using the ‘finalfit’, ‘survival’, and ‘cmprsk’ packages.^26^ The reporting of this study conforms to the STROBE statement.^28^

## Results

### Participants

At the LBC1936 study baseline, 1091 participants were initially recruited (49.8% female, mean [SD] age 69.5 [0.8] years). However, as the variables necessary to derive MCR were first measured at the six-year follow-up time point (wave 3), this became the baseline for our study (*n* = 697). We excluded one participant who did not consent to medical data linkage, six participants who developed dementia before wave 3, three participants who developed dementia less than one year after their wave 3 assessment, and seven participants missing data in one or more MCR criteria. A final total of 680 participants (48.3% female, mean [SD] age 76.2 [0.2] years) were included in our sample, giving a participation rate of eligible persons of 98% (680/697). The most common reasons for dropout in the LBC1936 are death, chronic incapacity, and permanent withdrawal.^16^ Figure 1 illustrates the participant flow and reasons for non-participation in this study.

**Figure 1.**
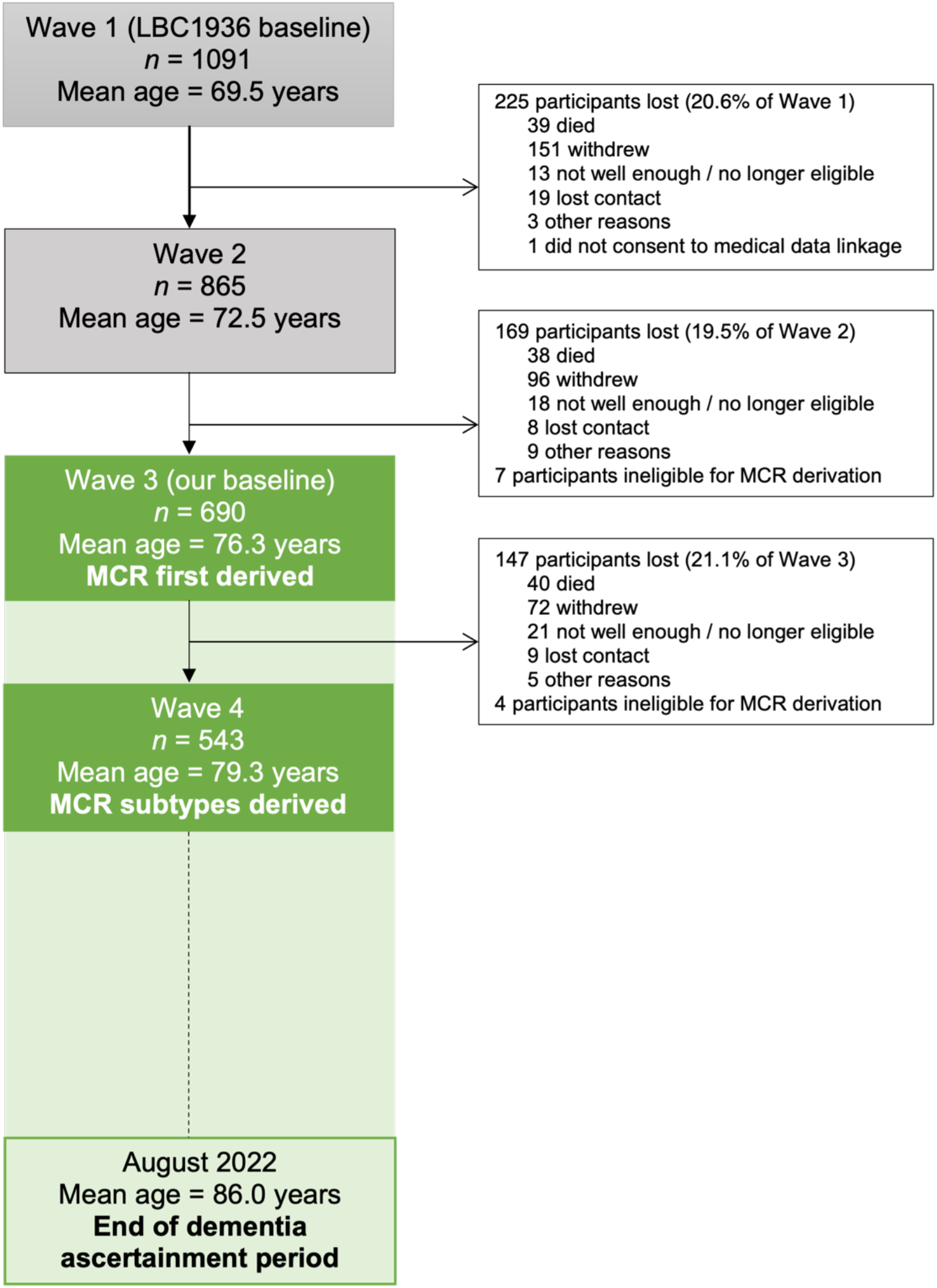
Flow chart of participants Note: MCR, Motoric Cognitive Risk; LBC1936, the Lothian Birth Cohort 1936. Dementia was ascertained in LBC1936 from wave 2 until August 2022, using medical data linkage. Therefore, all participants included in our baseline (wave 3) have been assessed for dementia. Green shading illustrates our study period. Waves 5 and 6 have now been completed but we did not require data from them (as we used medical data linkage), so have been excluded from the figure for clarity.

After 10 years of follow-up, 11.6% (*n* = 79/680) of the total cohort had developed dementia. MCR prevalence at wave 3 was 5.6% (95% CI 4.0–7.6; *n* = 38/680). Table 1 presents the characteristics of the study participants, comparing individuals who developed dementia with those who did not. MCR at baseline is a significant risk factor for developing dementia, as are fewer years of education and higher depression symptoms. There are no other significant differences in any demographic, socioeconomic, lifestyle, medical history, or physical or mental measures.

**Table 1.** Characteristics of study participants

|  |  | **Dementia** | **No Dementia** | **p** |
| --- | --- | --- | --- | --- |
| **MCR** | MCR | 9 (11.4) | 29 (4.8) | 0.032 |
|  | No MCR | 70 (88.6) | 572 (95.2) |  |
| **Age, years** | Mean (SD) | 76.2 (0.7) | 76.2 (0.7) | 0.259 |
| **Sex** | Female | 37 (46.2) | 293 (48.5) | 0.722 |
|  | Male | 43 (53.8) | 311 (51.5) |  |
| **Education, years** | Mean (SD) | 10.6 (1.0) | 10.8 (1.2) | 0.023 |
| **Occupational class** | Manual | 17 (21.8) | 118 (19.8) | 0.654 |
|  | Non-manual | 61 (78.2) | 479 (80.2) |  |
| **Physical activity level** | Low | 30 (37.5) | 179 (29.6) | 0.312 |
|  | Moderate | 34 (42.5) | 304 (50.3) |  |
|  | High | 14 (17.5) | 110 (18.2) |  |
| **Smoking history** | Current | 6 (7.5) | 38 (6.3) | 0.751 |
|  | Ex-smoker | 31 (38.8) | 255 (42.2) |  |
|  | Never | 43 (53.8) | 311 (51.5) |  |
| **Depression, HADS-D** | Mean (SD) | 3.4 (2.6) | 2.8 (2.2) | 0.035 |
| **Multimorbidity index** | Mean (SD) | 2.1 (1.3) | 2.3 (1.3) | 0.205 |
| **BMI, kg/m2** | Mean (SD) | 27.4 (3.8) | 27.8 (4.6) | 0.481 |

Note: MCR, Motoric Cognitive Risk; p, p-value; SD, Standard Deviation; HADS-D, Hospital Anxiety and Depression Scale - Depression; BMI, Body Mass Index; kg/m2, kilograms per metre squared.

### Main results

In older adults (average age of 76 years [SD 0.2]), the presence of MCR more than doubled the risk of incident dementia over the following 10 years. This finding was consistent across the basic model (aHR 2.83, 95% CI 1.41 to 5.67, p=0.003), the fully adjusted Cox regression model (aHR 2.45, 1.15 to 5.22, p=0.020), and the Fine-Gray competing risk model (aHR 2.34, 1.14 to 4.78, p=0.020). As expected, dementia was significantly associated with fewer formal years of education (p=0.023) and higher mean depressive symptoms (p=0.035). There was no significant difference in average ages between those with and without dementia.

The relationship over time between MCR and incident dementia is illustrated in Figure 2, with an accompanying risk table.

**Figure 2.**
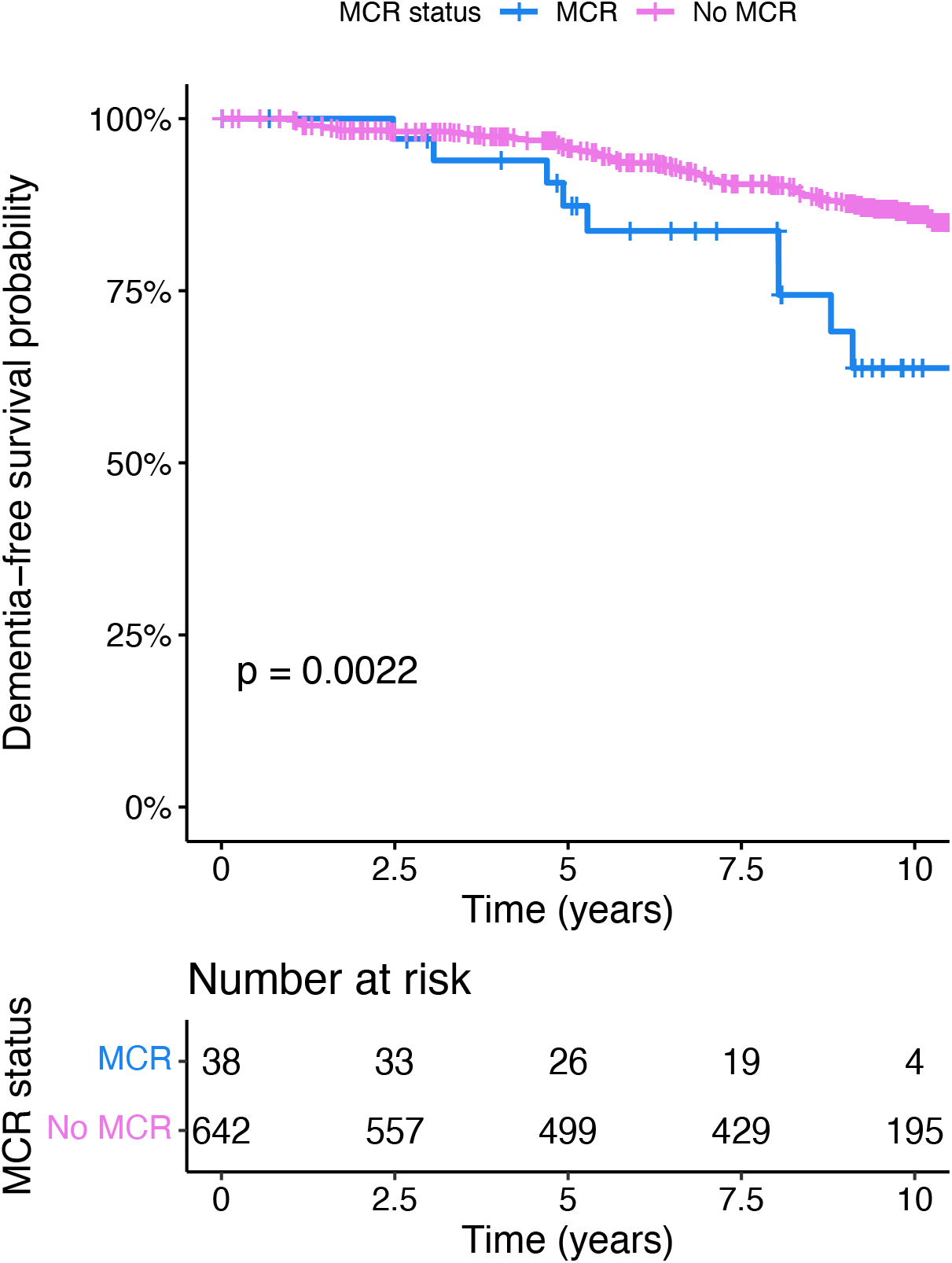
Kaplan-Meier survival curve for MCR and incident dementia over time, with accompanying risk table. Note: MCR; Motoric Cognitive Risk. The p-value is from a log-rank test that compared the cumulative survival rates between those with and without MCR.

Table 2 presents the results of unadjusted and adjusted Cox-proportional regression models and an adjusted Fine-Gray competing risk model. Dementia is the dependent variable, and MCR is the explanatory variable of interest. Potential confounders included in the adjusted models are presented in the table for completeness.

**Table 2.** Risk of incident dementia with Motoric Cognitive Risk syndrome

|  |  | N (%) | HR (DSS CPH unadjusted) | HR (DSS CPH adjusted) | HR (competing risks adjusted) |
| --- | --- | --- | --- | --- | --- |
| Dementia |  | 79 (11.6) |  |  |  |
| MCR | MCR | 38 (5.6) | 2.83 (1.41-5.67, p=0.003) | 2.45 (1.15-5.22, p=0.020) | 2.34 (1.14-4.78, p=0.020) |
|  | No MCR | 642 (94.4) | - | - | - |
| Age, years | Mean (SD) | 76.2 (0.7) | 1.14 (0.81-1.62, p=0.454) | 0.97 (0.66-1.42, p=0.872) | 0.91 (0.62-1.33, p=0.610) |
| Sex | Female | 330 (48.2) | - | - | - |
|  | Male | 354 (51.8) | 1.23 (0.79-1.91, p=0.358) | 1.31 (0.79-2.17, p=0.299) | 1.22 (0.73-2.03, p=0.450) |
| Education, years | Mean (SD) | 10.8 (1.1) | 0.78 (0.63-0.96, p=0.017) | 0.73 (0.58-0.93, p=0.011) | 0.73 (0.57-0.93, p=0.011) |
| Occupational class | Non-manual | 540 (80.0) | - | - | - |
|  | Manual | 135 (20.0) | 1.30 (0.76-2.22, p=0.342) | 0.76 (0.39-1.47, p=0.418) | 0.74 (0.39-1.40, p=0.350) |
| Multimorbidity index | Mean (SD) | 2.3 (1.3) | 0.95 (0.79-1.13, p=0.568) | 0.91 (0.75-1.10, p=0.335) | 0.86 (0.71-1.03, p=0.110) |
| Depression, HADS-D | Mean (SD) | 2.8 (2.3) | 1.15 (1.05-1.25, p=0.002) | 1.11 (1.01-1.21, p=0.036) | 1.10 (1.00-1.21, p=0.039) |
| BMI, kg/m2 | Mean (SD) | 27.7 (4.5) | 0.98 (0.93-1.04, p=0.532) | 0.96 (0.91-1.02, p=0.152) | 0.96 (0.92-1.01, p=0.150) |
| Physical activity level | High | 124 (18.5) | - | - | - |
|  | Low | 209 (31.1) | 1.40 (0.74-2.65, p=0.295) | 1.19 (0.60-2.37, p=0.621) | 1.22 (0.62-2.43, p=0.570) |
|  | Moderate | 338 (50.4) | 0.89 (0.48-1.65, p=0.705) | 0.82 (0.43-1.55, p=0.543) | 0.82 (0.43-1.55, p=0.530) |
| Smoking history | Never | 354 (51.8) | - | - | - |
|  | Current | 44 (6.4) | 1.80 (0.77-4.24, p=0.178) | 1.27 (0.50-3.23, p=0.614) | 0.82 (0.32-2.10, p=0.680) |
|  | Ex-smoker | 286 (41.8) | 0.96 (0.60-1.52, p=0.862) | 1.02 (0.62-1.67, p=0.945) | 0.95 (0.60-1.53, p=0.850) |

Note: N, total number; HR, Hazard Ratio; DSS, Disease-Specific Status; CPH Cox Proportional Hazard; p, p-value; SD, Standard Deviation; HADS-D, Hospital Anxiety and Depression Scale - Depression; BMI, Body Mass Index; kg/m2, kilograms per metre squared.

### Secondary analysis

As a secondary analysis, we followed the trajectories of the individuals with and without MCR over the three years from wave 3 (our baseline) to wave 4. This identified the MCR subgroups: Stable MCR (still have MCR; n=5), Transient Improved MCR (MCR at baseline but no MCR three years later due to an improvement – no longer a slow walker or no longer reported cognitive complaint), Transient Impaired MCR (MCR at baseline but no MCR three years later due to a deterioration – no longer functionally independent), and New MCR (developed MCR; n=22). We defined a fourth subgroup of those who never developed MCR at any time, Never MCR (n=483). For clarity, the classification period for transitioning between MCR states was the three years between wave 3 (baseline) and wave 4, while the classification period for transitioning from MCR state to Dementia was a maximum of 10 years – from baseline until the end of the LBC1936 dementia ascertainment period (August 2022).^12^ Of note, 15 participants with MCR at wave 3 but not wave 4 had improved (Transient Improved MCR). 13 of these participants were no longer classed as slow walkers and three no longer had a subjective cognitive complaint (one participant improved on both measures; Appendix 3.1). Three participants with MCR at wave 3 but not wave 4 had deteriorated (Transient Impaired MCR) as they were no longer classified as functionally independent (one of the four MCR criteria; Appendix 3.2). The sample sizes of these MCR subgroup are small so these findings should be interpreted with caution.

Appendix 3 compares the characteristics of individuals with and without each MCR subgroup classification. These tables are in the appendices as most subgroups are too small for meaningful interpretation. However, individuals in the largest subgroup, the Never MCR group (n=483), were significantly more likely to be younger (p<0.001), from a non-manual occupational background (p=0.002), have less depressive symptoms (p=0.016) and less likely to be sedentary (0.008), when compared with individuals who had MCR at any stage. Interestingly, over half of the Never MCR group still reported cognitive complaints at some stage, but less than one in 10 were classed as slow walkers at some stage.

The MCR transition pathways are illustrated in Figure 3. The thickness of the arrows in the illustration represents the proportion of participants transitioning from each starting state.

**Figure 3.**
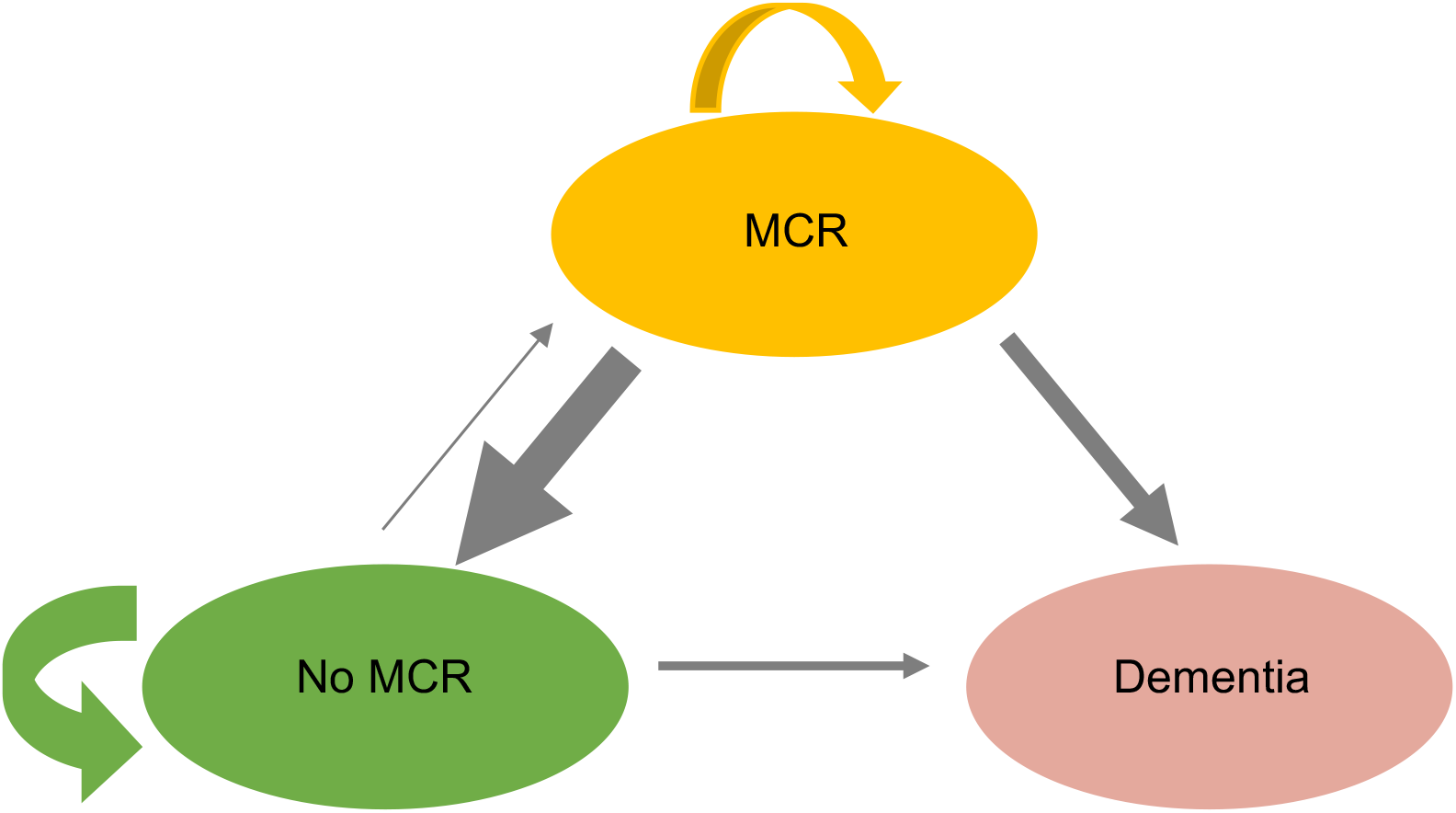
Transitions between MCR states over three years and dementia over ten years. Ovals specify possible states. Arrows specify possible transitions between states. Arrow thickness represents the proportion of each starting state transitioning to a different state. Transition arrows between No MCR and MCR (and vice-versa) states represent occurrences between baseline (wave 3) and three-year follow-up (wave 4). Follow-up for the dementia outcome was over a maximum of 10 years.

**Table 3.** Details of each Motoric Cognitive Risk transition state

| Transition label | Pathway | N | % |
| --- | --- | --- | --- |
| **Never MCR** | No MCR to No MCR | 483/642 | 75.2 |
| **New MCR** | No MCR to MCR | 22/642 | 3.4 |
| **Stable MCR** | MCR to MCR | 5/38 | 13.2 |
| **Transient Improved MCR\*** | MCR to No MCR | 15/38 | 39.5 |
| **Transient Impaired MCR^** | MCR to No MCR | 3/38 | 7.9 |
| **MCR Dementia** | MCR to Dementia | 9/38 | 23.7 |
| **No MCR Dementia** | No MCR to Dementia | 70/642 | 10.9 |

Note: N, Total number; %, percentage of total number; MCR, Motoric Cognitive Risk.
\* Transient Improved MCR subgroup participants were either no longer classed as slow walkers or no longer reported subjective cognitive complaints (or both).
^ Transient Impaired MCR subgroup participants were no longer functionally independent (one of the MCR criteria).

Figure 4 illustrates Kaplan-Meier estimates of dementia-free survival differences between the MCR subgroups and includes a number-at-risk table. The size of some groups, especially Transient Impaired MCR and Stable MCR, are small, so should be interpreted with caution.

**Figure 4.**
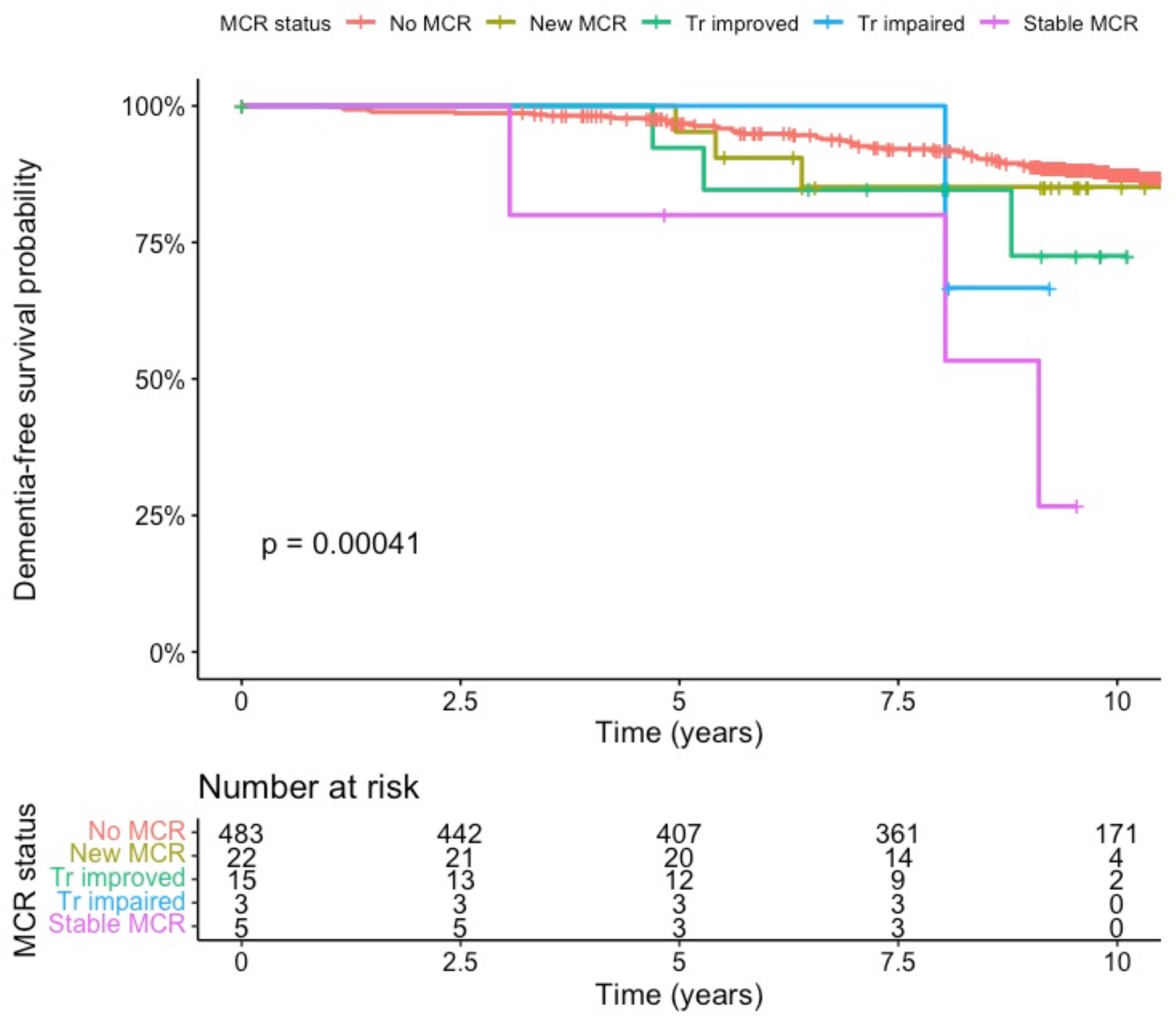
Kaplan-Meier survival curve for MCR subgroups and incident dementia over time, with accompanying risk table. Note: MCR, Motoric Cognitive Risk. Tr, Transient. The p-value is from a log-rank test of the trend of the cumulative survival rates. Subgroups were defined by following the trajectories of participants between baseline (wave 3) and three-year follow-up (wave 4).

Table 4 presents our analysis of the MCR subgroups and the risk of dementia. We have included a caveat that they should be interpreted with caution due to the sample size. However, it is interesting to note the increasing hazard ratio for incident dementia when moving through the MCR subgroups of New MCR (aHR 1.08, 95% CI 0.29-4.05, p=0.910), Transient Improved MCR (aHR 1.83 95% CI 0.53-6.32, p=0.340), Stable MCR (aHR 4.38, 95% CI 1.43-13.44, p=0.010), and finally Transient Impaired MCR (aHR 8.15 95% CI 1.37-48.60, p=0.021).

**Table 4.** Motoric Cognitive Risk subgroups and risk of incident dementia

| MCR subgroup | Eligible sample, N (%) | Incident dementia, N (%) | CPH | CPH | CRR |
| --- | --- | --- | --- | --- | --- |
|  |  |  | Unadjusted HR (95% CI), p-value | Adjusted HR (95% CI), p-value | Adjusted HR (95% CI), p-value |
| **Never MCR** | 483 (89.0) | 50 (10.3) | 0.40 (0.22-0.76, $p=0.005$) | 0.48 (0.23-0.99, $p=0.046$) | 0.52 (0.25-1.09, $p=0.084$) |
| **New MCR** | 22 (4.0) | 3 (13.6) | 1.24 (0.39-3.94, $p=0.720$) | 1.02 (0.31-3.41, $p=0.971$) | 1.08 (0.29-4.05, $p=0.910$) |
| **Transient Improved** | 15 (2.2) | 3 (20) | 1.97 (0.62-6.26, $p=0.249$) | 1.76 (0.54-5.78, $p=0.348$) | 1.83 (0.53-6.32, $p=0.340$) |
| **Stable MCR** | 5 (0.7) | 3 (60) | 6.70 (2.11-21.28, $p=0.001$) | 3.53 (0.92-13.56, $p=0.066$) | 4.38 (1.43-13.44, $p=0.010$) |
| **Transient Impaired** | 3 (0.4) | 1 (33) | 2.78 (0.39-20.04, $p=0.310$) | 6.57 (0.78-55.19, $p=0.083$) | 8.15 (1.37-48.60, $p=0.021$) |

Note: CPH, Cox-Proportional Hazard; HR, Hazard Ratio; CI, Confidence Interval; CRR, Competing Risk Regression; MCR, Motoric Cognitive Risk; p, P-value. The percentage in the incident dementia column is in relation to the number in the subgroup. These results should be interpreted with caution as the size of some subgroups, especially Stable MCR, is very small.

## Discussion

### Key results

In this community-based longitudinal study, we have demonstrated that MCR, the co-occurrence of slow gait and cognitive complaints, is associated with a greater than two-fold increase in risk for incident dementia. This is similar to previous findings and reinforces the potential clinical utility of MCR, within a Scottish context.^6–8,29,30^ Our finding remained robust after accounting for death in a competing risk regression. We believe ours is the first MCR study to use a competing risk approach to time-to-event analysis with dementia as the outcome. This is a strength of our work as it is crucial to account for the competing risk of death precluding dementia as our primary outcome of interest as our participants were, on average, 76 years at baseline and were followed up for up to 10 years. That the effect size (aHR) is only slightly reduced after accounting for competing risk, in comparison to the Cox-proportional hazard model, is possibly partly due to the healthy nature of the LBC1936 participants. Our study’s baseline was wave 3 of the LBC1936 study. Many participants who dropped out of LBC1936 by wave 3 (our baseline) were those who died or had poorer health.^16^ Regardless, it is likely that our estimates are more accurate than MCR studies using traditional survival analysis methods alone, particularly in studies with an older population.^25,31^

Of the potential confounders included, dementia was significantly associated with fewer formal years of education and higher mean depressive symptoms. Both have been consistently associated with an increased risk of incident MCR in the literature, with a recent meta-analysis reporting the following associations between MCR and education (8 studies; OR 2.04, 95% CI 1.28 to 3.25) and depression (17 studies; OR 2.19, 95% CI 1.65 to 2.9). We maintained the depression measure (HADS-D) as a continuous measure in our analysis, given it is a symptom rather than diagnostic scale. Our study found no difference in the average ages between those with and without dementia. This is likely due to the very narrow age spread amongst the LBC1936 participants (SD 0.7 years), all of whom were born in 1936.^17^

Our secondary analysis illustrates the heterogeneous nature of MCR progression and highlights that not all older adults with MCR will follow a similar path. It is true that some of our secondary analysis results are based on small numbers and are of an explorative nature. Nonetheless, we found that being classed in either the New MCR or Transient Improved MCR subgroups did not significantly increase the risk of subsequent incident dementia. However, being classed as Stable MCR increased the risk of dementia four-fold and Transient Impaired MCR eight-fold, even after accounting for competing risks and adjusting for potential confounders. Crucially, though, only five participants were classed as having Stable MCR and three as having Transient Impaired MCR, so this finding is non-conclusive. Our finding that only some subgroups of MCR are associated with an increased incident dementia risk is in contrast to a recent paper which found that all MCR subgroups predicted incident dementia.^14^ That study, however, grouped transient impaired and improved individuals together, potentially diluting the effect of both.^14^ Further work exploring the important aspect of MCR trajectories, preferably using a large MCR consortium of cohorts, is merited, as both studies examining it to date have limited sample sizes. Ideally, cohorts with imaging data should be included to allow for the exploration of the biological mechanisms underpinning any differences between MCR subgroups, given their different risk profiles for dementia.

The Transient Improved MCR group consisted of 15 participants with MCR at wave 3 who were classed as No MCR at wave 4. Interestingly, at wave 4, only three of these participants no longer had a subjective cognitive complaint, while 13 participants were no longer classed as slow walkers. One critique levelled at using the subjective cognitive complaint measure is that people may report a cognitive complaint one day but not the next, thus rendering it unreliable.^32^ Our analysis, albeit on a small sample and therefore not conclusive, indicates this is unlikely the case in our cohort. That some individuals with MCR at baseline progressed beyond having MCR by way of losing functional independence is in keeping with a previously reported association between MCR and incident disability.^8,33^

Participants who never developed MCR at any stage (Never MCR) were the largest subgroup (n = 483). Individuals in this group were significantly more likely to be younger, from a non-manual occupational background, have fewer depressive symptoms, and be more physically active when compared with individuals who had MCR at any stage (Appendix 3.5). Of note, over half of the Never MCR group reported cognitive complaints at some stage. This seemingly high rate of subjective cognitive complaints is, in fact, lower than the rates commonly reported in older adults, where up to 88% of older adults in community settings have complained of memory problems.^34^ Less than one in 10 of the Never MCR subgroup were classed as slow walkers at any stage, indicating that slow gait has a good differential utility, complementing the more common subjective cognitive complaint measure.

### Context within the literature

It is difficult to place the MCR trajectory analysis component of our study in context in the literature beyond the already referenced only other study to analyse MCR trajectories.^14^ However, Mild Cognitive Impairment (MCI) is a predementia syndrome that has been studied more and over a longer period, including analyses of MCI trajectories.^35^ As MCI and MCR are both predementia syndromes sharing similar operational constructs, it is no surprise that individuals with MCR follow similar trajectories to those reported in the MCI literature.^35,36^ A recent study of the bidirectional transitions of MCI (reversion and progression) in 6,651 participants used a multistate modelling approach to estimate instantaneous transition intensity between the states and transition probabilities from one state to another at any given time during follow-up.^36^ The authors found that post-reversion participants remained at an increased risk of progression to MCI or dementia over the longer term and experience recurrent reversions.^36^ If the LBC1936 were a larger dataset, we would have liked to use multistate modelling approaches in our study to analyse if the same were true of our data. Figure 3 is a typical image used in multistate modelling approaches to illustrate the state structure and possible transitions, adapted for our study to account for the smaller sample size.

### Limitations

A further limitation of our data includes the risk of attrition bias. Despite the best efforts of the LBC1936 research team to minimise the dropout rate, it is approximately 20% between waves. This resulted in a 37% reduction in participants over the six years between wave 1 and wave 3, when MCR was first derived. This dropout rate, although substantial, remains within the acceptable limit suggested by international quality assessment bodies.^37^ Only 17 of the 697 (2.4%) available participants were excluded, for reasons detailed in Figure 1. This high participation rate helps alleviate any selection bias concerns. The robust dementia outcome now available in LBC1936 uses medical data linkage for follow-up, which all but negates any risk of attrition bias for that outcome. Nevertheless, our sample size remains small. Our findings would engender more confidence if replicated in a larger cohort or in a cohort with a higher prevalence of MCR.

### Implications and generalisability

Our findings have several implications. First, if the association between MCR and incident dementia reflects a causal link, health and social policy measures which target the modifiable risk factors of MCR in early to mid-life might reduce the numbers of individuals transitioning to MCR and then to dementia. Meta-analyses of risk factors for MCR have identified several targets which are also associated with increased dementia risk.^5,11,21^ These would be a good starting point and include: diabetes (21 studies; OR 1.50, 95%CI 1.37 to 1.64), hypertension (21 studies; OR 1.20, 95% CI 1.08 to 1.33), stroke (16 studies; OR 2.03, 95% CI 1.70 to 2.42), heart disease (7 studies; OR 1.45, 95% CI 1.13 to 1.86), coronary artery disease (5 studies; OR 1.49, 95% CI1.16 to 1.91), smoking (13 studies; OR 1.28, 95% CI 1.04 to 1.58), and obesity (12 studies; OR 1.34, 95% CI 1.13 to 1.59).^21^ Second, given the clinical utility of MCR and ease of diagnosis, consideration should be given to incorporating its use into brain health clinics in Scotland. This would likely entail adding a brief walking speed assessment during brain health clinics, as subjective cognitive complaints and functional ability are already routinely assessed. However, to determine whether an individual is a slow walker, it is imperative to first determine robust national age- and sex-matched slow gait speed cut-offs. This is an important next step. Third, our findings that higher depressive symptoms are a risk factor for dementia reinforce previous research which linked depression to both MCR and dementia.^20,21,30,38,39^ As a modifiable risk factor, depression could be a target for any future trials assessing if preventing MCR leads to a reduction in incident dementia.

When applying our findings to other populations, it is important to note that the LBC1936 is not a nationally representative sample. The participants in LBC1936 have a higher average number of years of education and better general physical fitness than the Scottish population.^16^ Participants are also all white.^16^

## Conclusion

In conclusion, our prospective study provides further support that the clinical syndrome, MCR, identifies older individuals at high risk for transitioning to dementia. Identifying MCR is recommended for early detection and instituting preventative measures for reducing the risk of dementia. Our secondary analysis illustrates the heterogeneous nature of MCR progression. MCR subtype status influenced its association with incident dementia, with the Stable MCR and Transient Impaired MCR subgroups identifying high-risk individuals, while the Transient Improved MCR and New MCR subgroups did not. This subtyping data is preliminary, and it will be important that future work confirms it in larger datasets or, preferably, in multiple cohorts.

## Data Availability

All data produced in the present study are available upon reasonable request to the authors. All data is available from https://www.ed.ac.uk/lothian-birth-cohorts/data-access-collaboration

## Acknowledgements

The authors are very grateful to the LBC1936 participants, past and present. Their commitment and contributions have made this work and many other important studies possible. Dr Mullin is very grateful for the support of his funders, supervisors, and academic mentors. For the purpose of open access, the authors have applied a CC-BY public copyright licence to any Author Accepted Manuscript version arising from this submission.

## Appendix 1

Illustration of the missing values map for each covariate.

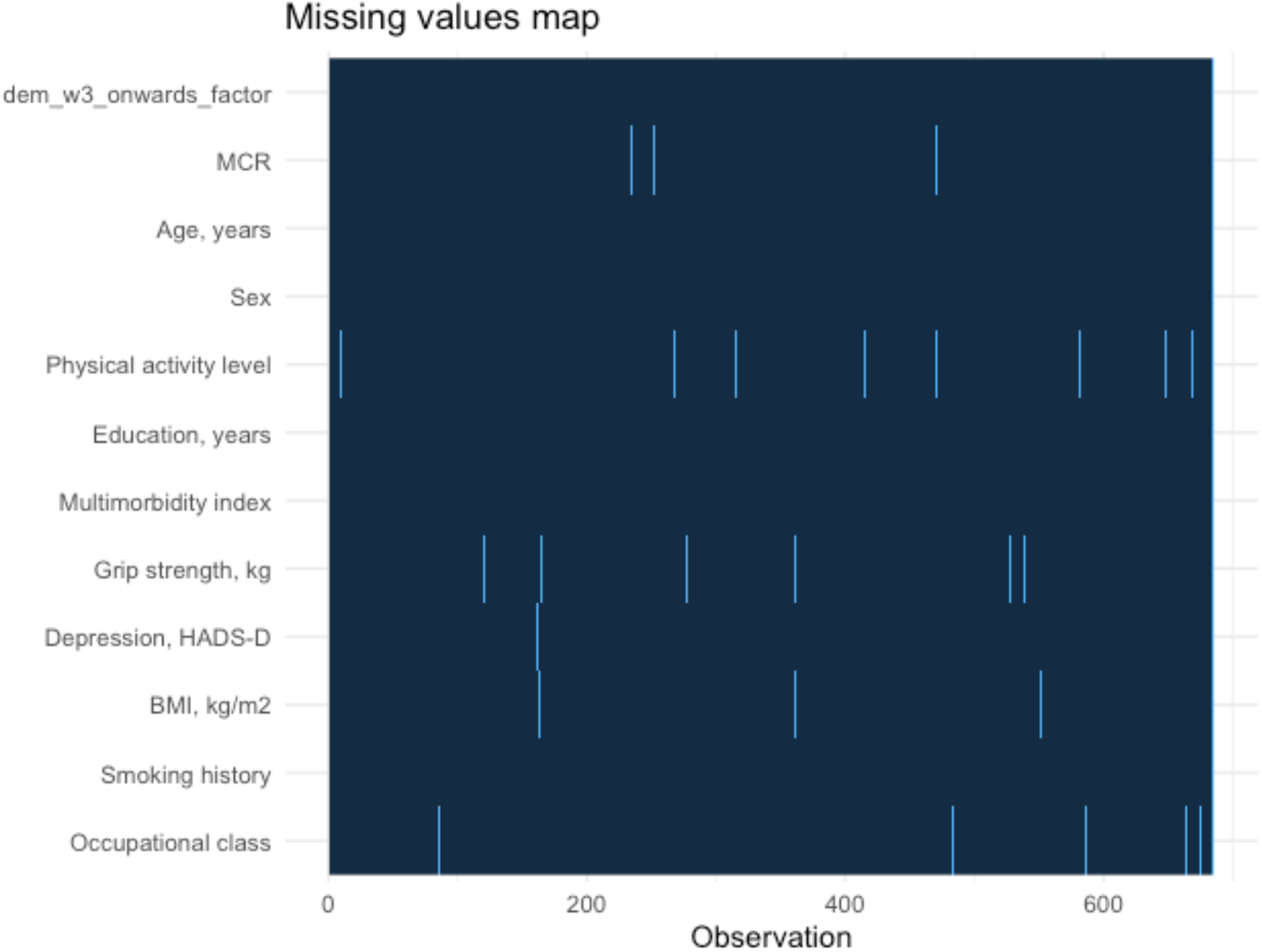

Note: each blue line indicates a missing variable for one participant. There is no obvious pattern to any missingness, indicating data is more likely to be missing at random.

## Appendix 2

Categorisation of the physical activity level variable

The original self-reported physical activity levels codes in LBC1936 are:

1 = moving only in connection with necessary household chores;
2 = walking or other outdoor activities 1–2 times per week;
3 = walking or other outdoor activities several times per week;
4 = exercising 1–2 times per week to the point of perspiring and heavy breathing;
5 = exercising several times per week to the point of perspiring and heavy breathing;
6 = keep fit/heavy exercise or competitive sport several times weekly.

To improve the distribution and reduce the spread of data for our model, we categorized self-reported physical activity levels 1 and 2 into “Low”, 3 and 4 into “Medium”, and 5 and 6 into “High”.

## Appendix 3

MCR subgroups – demographics tables and time-to-event models for each subgroup

**Appendix 3.1:** Transient improved MCR subgroup demographics

| label | levels | No Transient improved | Transient improved | Total | p |
| --- | --- | --- | --- | --- | --- |
| **Total N (%)** |  | 652 (97.8) | 15 (2.2) | 667 |  |
| **ageyears_w4** | Mean (SD) | 79.3 (0.6) | 79.7 (0.5) | 79.3 (0.6) | 0.004 |
| **Sex.factor** | Female | 314 (48.2) | 5 (33.3) | 319 (47.8) | 0.303 |
|  | Male | 338 (51.8) | 10 (66.7) | 348 (52.2) |  |
|  | (Missing) | 0 (0.0) | 0 (0.0) | 0 (0.0) |  |
| **yrsedu_w1** | Mean (SD) | 10.8 (1.1) | 10.5 (1.0) | 10.8 (1.1) | 0.301 |
| **Social.factor.collapsed** | Non-manual | 518 (79.4) | 10 (66.7) | 528 (79.2) | 0.191 |
|  | Manual | 125 (19.2) | 5 (33.3) | 130 (19.5) |  |
|  | (Missing) | 9 (1.4) | 0 (0.0) | 9 (1.3) |  |
| **vasc_comorbidity.factor_w3** | None | 204 (31.3) | 4 (26.7) | 208 (31.2) | 0.527 |
|  | Moderate | 401 (61.5) | 9 (60.0) | 410 (61.5) |  |
|  | High | 47 (7.2) | 2 (13.3) | 49 (7.3) |  |
| **HADS_D_w4** | Mean (SD) | 3.0 (2.3) | 3.3 (2.6) | 3.0 (2.3) | 0.582 |
| **bmi_w4** | Mean (SD) | 27.3 (4.5) | 27.9 (4.6) | 27.3 (4.5) | 0.619 |
| **phyactiv.factor_w3** | High | 119 (18.3) | 2 (13.3) | 121 (18.1) | 0.778 |
|  | Low | 195 (29.9) | 6 (40.0) | 201 (30.1) |  |
|  | Moderate | 326 (50.0) | 7 (46.7) | 333 (49.9) |  |
|  | (Missing) | 12 (1.8) | 0 (0.0) | 12 (1.8) |  |
| **Smoking.factor.w4** | Current | 20 (3.9) |  | 20 (3.8) | 0.689 |
|  | Ex-smoker | 219 (42.4) | 5 (33.3) | 224 (42.1) |  |
|  | Never | 278 (53.8) | 10 (66.7) | 288 (54.1) |  |
| **Memprob.factor.w4** | Memory prob | 292 (58.3) | 11 (78.6) | 303 (58.8) | 0.171 |
|  | No memory prob | 209 (41.7) | 3 (21.4) | 212 (41.2) |  |
| **Slow.factor.w4** | Not slow | 434 (85.8) | 13 (86.7) | 447 (85.8) | 1.000 |
|  | Slow | 72 (14.2) | 2 (13.3) | 74 (14.2) |  |

**Appendix 3.2:** Transient impaired MCR subgroup demographics

| label | levels | No Transient<br>impaired | Transient<br>impaired | Total | p |
| --- | --- | --- | --- | --- | --- |
| **Total N (%)** |  | 664 (99.6) | 3 (0.4) | 667 |  |
| **ageyears_w4** | Mean (SD) | 79.3 (0.6) | 79.8 (0.3) | 79.3 (0.6) | 0.144 |
| **Sex.factor** | Female | 318 (47.9) | 2 (66.7) | 320 (48.0) | 0.610 |
|  | Male | 346 (52.1) | 1 (33.3) | 347 (52.0) |  |
| **yrsedu_w1** | Mean (SD) | 10.8 (1.1) | 11.0 (0.0) | 10.8 (1.1) | <0.001 |
| **Social.factor.collapsed** | Manual | 127 (19.4) | 2 (100.0) | 129 (19.6) | 0.038 |
|  | Non-manual | 529 (80.6) |  | 529 (80.4) |  |
| **comorbidity_w4** | Mean (SD) | 1.9 (1.5) | 2.0 (1.0) | 1.9 (1.5) | 0.930 |
| **HADS_D_w4** | Mean (SD) | 2.9 (2.3) | 8.3 (6.1) | 3.0 (2.3) | 0.266 |
| **bmi_w4** | Mean (SD) | 27.4 (4.5) | 24.3 (1.5) | 27.3 (4.5) | 0.065 |
| **phyactiv.factor_w3** | Low | 199 (30.0) | 2 (66.7) | 201 (30.1) | 0.588 |
|  | Moderate | 332 (50.0) | 1 (33.3) | 333 (49.9) |  |
|  | High | 120 (18.1) | 0 (0.0) | 120 (18.0) |  |
|  | (Missing) | 13 (2.0) | 0 (0.0) | 13 (1.9) |  |
| **Smoking.factor.w4** | Current | 20 (3.8) | 1 (33.3) | 21 (3.9) | 0.060 |
|  | Ex-smoker | 223 (42.2) |  | 223 (41.9) |  |
|  | Never | 286 (54.1) | 2 (66.7) | 288 (54.1) |  |
| **Memprob.factor.w4** | Memory prob | 298 (58.2) | 3 (100.0) | 301 (58.4) | 0.270 |
|  | No memory prob | 214 (41.8) |  | 214 (41.6) |  |
| **Slow.factor.w4** | Not slow | 448 (86.3) |  | 448 (86.0) | 0.019 |
|  | Slow | 71 (13.7) | 2 (100.0) | 73 (14.0) |  |

**Appendix 3.3:** New MCR subgroup demographics

| label | levels | No New MCR | New MCR | Total | p |
| --- | --- | --- | --- | --- | --- |
| **Total N (%)** |  | 524 (96.0) | 22 (4.0) | 546 |  |
| **ageyears_w4** | Mean (SD) | 79.3 (0.6) | 79.6 (0.6) | 79.3 (0.6) | 0.022 |
| **Sex** | Female | 258 (49.2) | 11 (50.0) | 269 (49.3) | 1.000 |
|  | Male | 266 (50.8) | 11 (50.0) | 277 (50.7) |  |
|  | (Missing) | 0 (0.0) | 0 (0.0) | 0 (0.0) |  |
| **Education, years** | Mean (SD) | 10.9 (1.2) | 10.7 (1.2) | 10.9 (1.2) | 0.477 |
| **Occupational class** | Non-manual | 422 (80.5) | 17 (77.3) | 439 (80.4) | 0.574 |
|  | Manual | 93 (17.7) | 5 (22.7) | 98 (17.9) |  |
|  | (Missing) | 9 (1.7) | 0 (0.0) | 9 (1.6) |  |
| **comorbidity_w4** | Mean (SD) | 2.4 (1.3) | 2.7 (1.1) | 2.4 (1.3) | 0.181 |
| **HADS_D_w4** | Mean (SD) | 2.9 (2.3) | 3.8 (2.2) | 3.0 (2.3) | 0.096 |
| **bmi_w4** | Mean (SD) | 27.3 (4.5) | 28.9 (5.8) | 27.4 (4.5) | 0.204 |
| **Physical activity level** | High | 100 (19.1) | 7 (31.8) | 107 (19.6) | 0.140 |
|  | Low | 150 (28.6) | 8 (36.4) | 158 (28.9) |  |
|  | Moderate | 263 (50.2) | 7 (31.8) | 270 (49.5) |  |
|  | (Missing) | 11 (2.1) | 0 (0.0) | 11 (2.0) |  |
| **Smoking.factor.w4** | Current | 19 (3.7) | 2 (9.1) | 21 (3.9) | 0.226 |
|  | Ex-smoker | 218 (42.7) | 7 (31.8) | 225 (42.2) |  |
|  | Never | 274 (53.6) | 13 (59.1) | 287 (53.8) |  |
| **Memprob.factor.w4** | Memory prob | 280 (56.7) | 22 (100.0) | 302 (58.5) | <0.001 |
|  | No memory prob | 214 (43.3) |  | 214 (41.5) |  |
| **Slow.factor.w4** | Not slow | 448 (89.6) |  | 448 (85.8) | <0.001 |
|  | Slow | 52 (10.4) | 22 (100.0) | 74 (14.2) |  |

**Appendix 3.4:** Stable MCR Demographics Table

| label | levels | No Stable MCR | Stable MCR | p |
| --- | --- | --- | --- | --- |
| **ageyears_w4** | Mean (SD) | 79.3 (0.6) | 79.5 (0.5) | 0.533 |
| **Sex** | Female | 317 (47.8) | 4 (80.0) | 0.200 |
|  | Male | 346 (52.2) | 1 (20.0) |  |
|  | (Missing) | 0 (0.0) | 0 (0.0) |  |
| **Education, years** | Mean (SD) | 10.8 (1.1) | 10.4 (1.5) | 0.581 |
| **Occupational class** | Non-manual | 527 (79.5) | 3 (60.0) | 0.254 |
|  | Manual | 127 (19.2) | 2 (40.0) |  |
|  | (Missing) | 9 (1.4) | 0 (0.0) |  |
| **comorbidity_w4** | Mean (SD) | 1.9 (1.5) | 2.2 (1.3) | 0.679 |
| **HADS_D_w4** | Mean (SD) | 3.0 (2.3) | 4.2 (3.0) | 0.415 |
| **bmi_w4** | Mean (SD) | 27.4 (4.5) | 25.5 (4.2) | 0.386 |
| **Physical activity level** | High | 120 (18.1) | 0 (0.0) | 0.405 |
|  | Low | 198 (29.9) | 3 (60.0) |  |
|  | Moderate | 332 (50.1) | 2 (40.0) |  |
|  | (Missing) | 13 (2.0) | 0 (0.0) |  |
| **Smoking.factor.w4** | Current | 20 (3.8) | 1 (20.0) | 0.241 |
|  | Ex-smoker | 222 (42.0) | 2 (40.0) |  |
|  | Never | 286 (54.2) | 2 (40.0) |  |
| **Memprob.factor.w4** | Memory prob | 296 (58.0) | 5 (100.0) | 0.079 |
|  | No memory prob | 214 (42.0) |  |  |
| **Slow.factor.w4** | Not slow | 448 (86.8) |  | <0.001 |
|  | Slow | 68 (13.2) | 5 (100.0) |  |

**Appendix 3.5:**
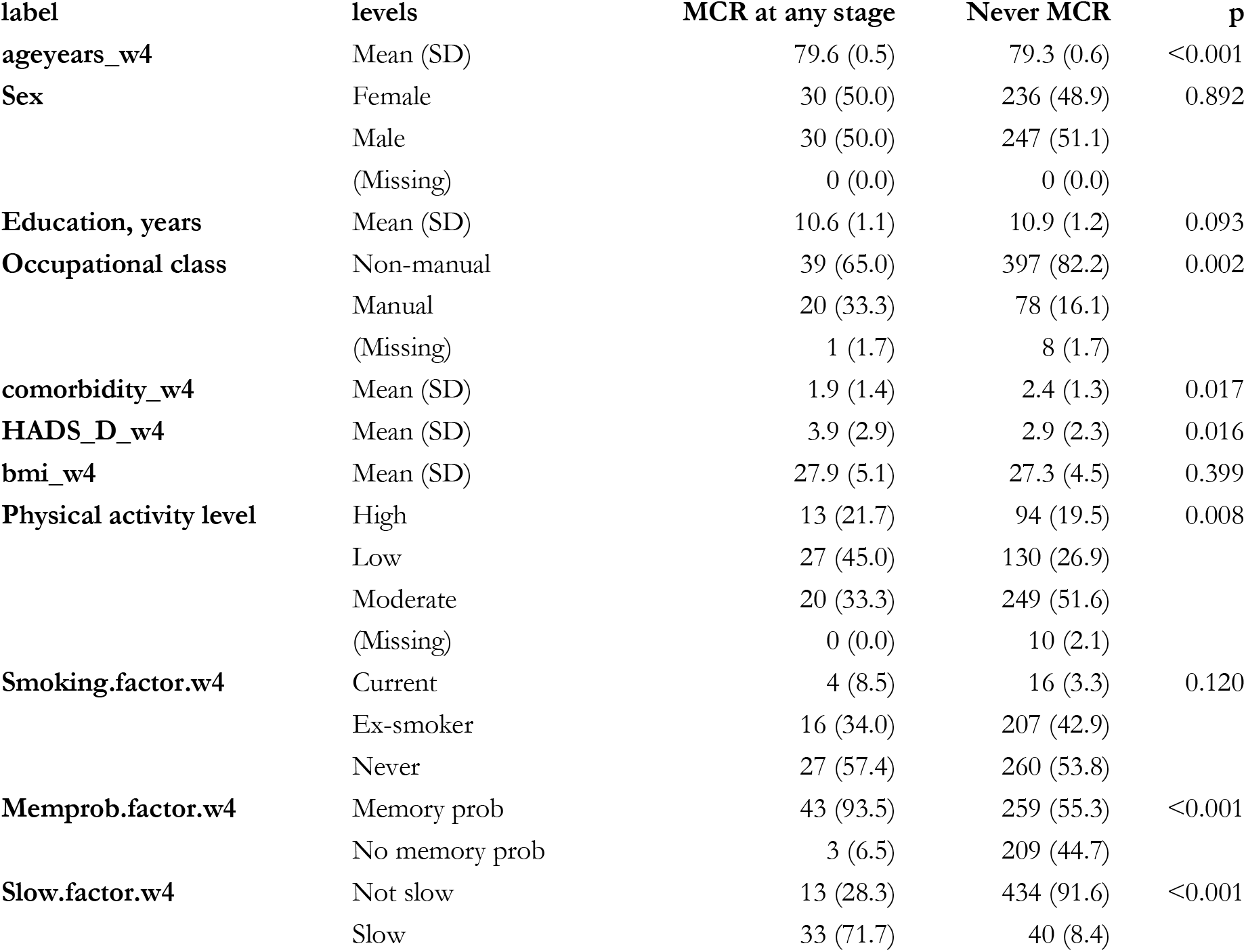
Never MCR Demographics Table

**Appendix 3.6:**
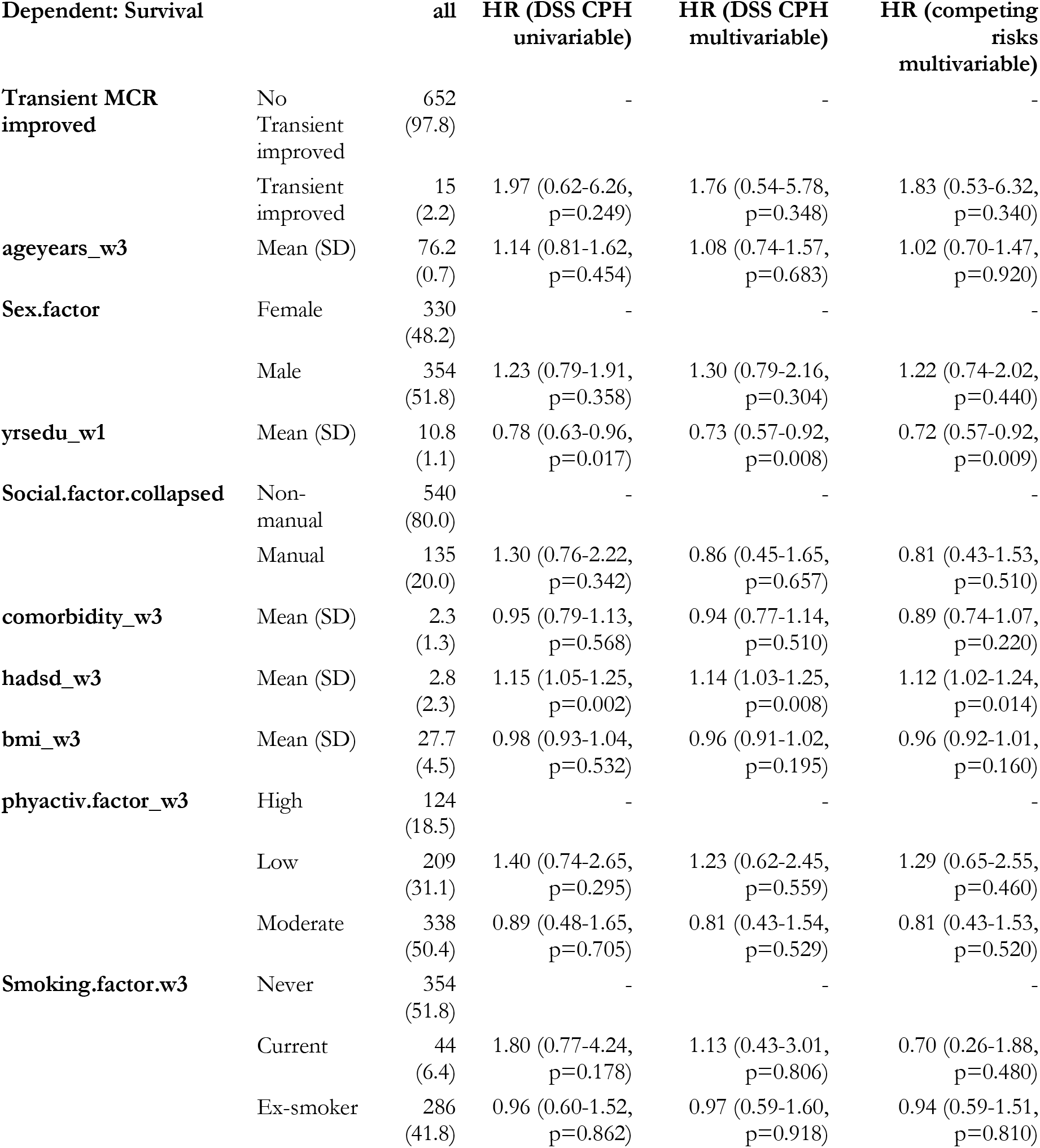
Transient Improved MCR time-to-event model

**Appendix 3.7:**
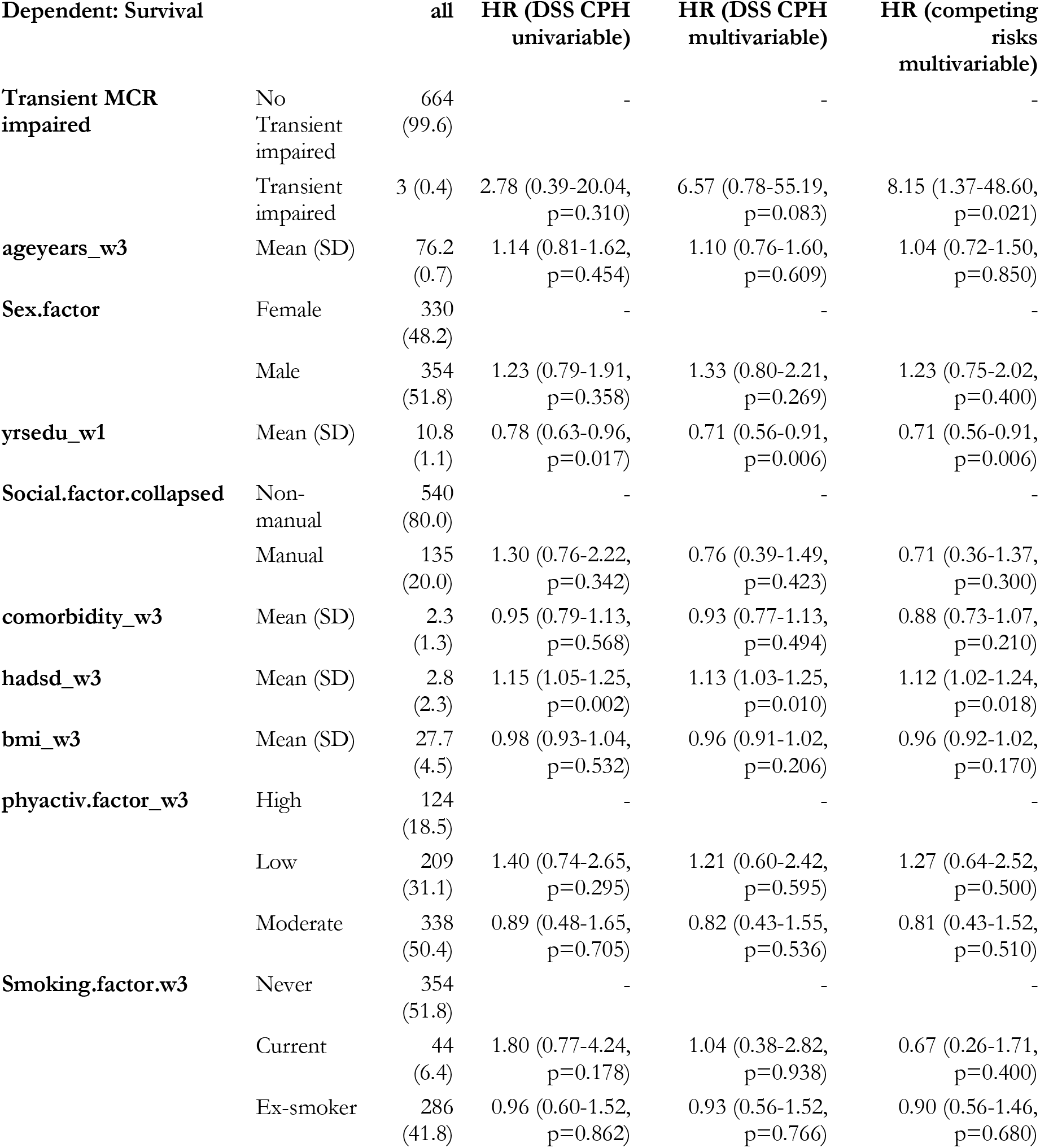
Transient impaired time-to-event model

**Appendix 3.8:** New MCR time-to-event model

| Dependent:<br>Survival |  | all | HR (DSS CPH<br>univariable) | HR (DSS CPH<br>multivariable) | HR (competing<br>risks multivariable) |
| --- | --- | --- | --- | --- | --- |
| New MCR | No New MCR | 524<br>(96.0) | - | - | - |
|  | New MCR | 22<br>(4.0) | 1.24 (0.39-3.94,<br>p=0.720) | 1.02 (0.31-3.41,<br>p=0.971) | 1.08 (0.29-4.05,<br>p=0.910) |
| Age, years | Mean (SD) | 76.2<br>(0.7) | 1.14 (0.81-1.62,<br>p=0.454) | 1.12 (0.73-1.71,<br>p=0.613) | 1.03 (0.66-1.60,<br>p=0.910) |
| Sex | Female | 330<br>(48.2) | - | - | - |
|  | Male | 354<br>(51.8) | 1.23 (0.79-1.91,<br>p=0.358) | 1.03 (0.58-1.83,<br>p=0.910) | 0.98 (0.55-1.76,<br>p=0.950) |
| Education, years | Mean (SD) | 10.8<br>(1.1) | 0.78 (0.63-0.96,<br>p=0.017) | 0.76 (0.58-0.98,<br>p=0.037) | 0.76 (0.59-0.98,<br>p=0.033) |
| Occupational<br>class | Non-manual | 540<br>(80.0) | - | - | - |
|  | Manual | 135<br>(20.0) | 1.30 (0.76-2.22,<br>p=0.342) | 1.17 (0.56-2.42,<br>p=0.678) | 1.14 (0.54-2.38,<br>p=0.730) |
| comorbidity_w3 | Mean (SD) | 2.3<br>(1.3) | 0.95 (0.79-1.13,<br>p=0.568) | 0.91 (0.73-1.13,<br>p=0.397) | 0.88 (0.73-1.07,<br>p=0.210) |
| Depression,<br>HADS-D | Mean (SD) | 2.8<br>(2.3) | 1.15 (1.05-1.25,<br>p=0.002) | 1.10 (0.99-1.23,<br>p=0.074) | 1.11 (0.99-1.24,<br>p=0.070) |
| BMI, kg/m2 | Mean (SD) | 27.7<br>(4.5) | 0.98 (0.93-1.04,<br>p=0.532) | 0.96 (0.90-1.03,<br>p=0.238) | 0.96 (0.91-1.02,<br>p=0.200) |
| Physical activity<br>level | High | 124<br>(18.5) | - | - | - |
|  | Low | 209<br>(31.1) | 1.40 (0.74-2.65,<br>p=0.295) | 1.26 (0.58-2.72,<br>p=0.562) | 1.23 (0.57-2.66,<br>p=0.590) |
|  | Moderate | 338<br>(50.4) | 0.89 (0.48-1.65,<br>p=0.705) | 0.82 (0.41-1.65,<br>p=0.582) | 0.84 (0.43-1.64,<br>p=0.600) |
| Smoking history | Never | 354<br>(51.8) | - | - | - |
|  | Current | 44<br>(6.4) | 1.80 (0.77-4.24,<br>p=0.178) | 1.75 (0.57-5.39,<br>p=0.328) | 1.55 (0.47-5.17,<br>p=0.470) |
|  | Ex-smoker | 286<br>(41.8) | 0.96 (0.60-1.52,<br>p=0.862) | 1.13 (0.65-1.95,<br>p=0.673) | 1.12 (0.66-1.91,<br>p=0.670) |

**Appendix 3.9:**
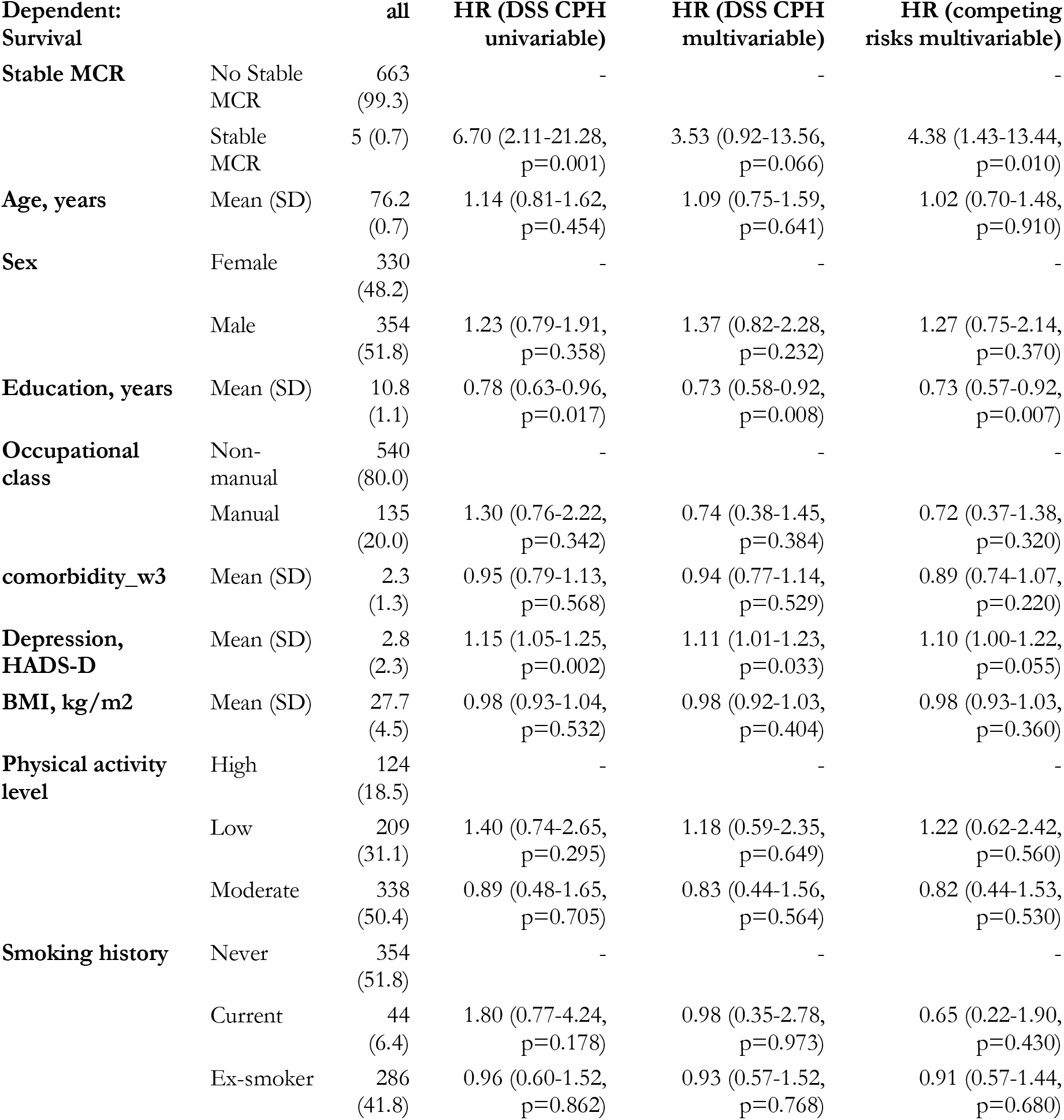
Stable MCR time-to-event model

**Appendix 3.10:** Never MCR time-to-event model

| Dependent:<br>Survival |  | all | HR (DSS CPH<br>univariable) | HR (DSS CPH<br>multivariable) | HR (competing<br>risks multivariable) |
| --- | --- | --- | --- | --- | --- |
| Never MCR | MCR at any stage | 60<br>(11.0) | - | - | - |
|  | Never MCR | 483<br>(89.0) | 0.40 (0.22-0.76,<br>p=0.005) | 0.48 (0.23-0.99,<br>p=0.046) | 0.52 (0.25-1.09,<br>p=0.084) |
| Age, years | Mean (SD) | 76.2<br>(0.7) | 1.14 (0.81-1.62,<br>p=0.454) | 1.03 (0.67-1.60,<br>p=0.885) | 0.95 (0.60-1.51,<br>p=0.830) |
| Sex | Female | 330<br>(48.2) | - | - | - |
|  | Male | 354<br>(51.8) | 1.23 (0.79-1.91,<br>p=0.358) | 1.08 (0.61-1.93,<br>p=0.786) | 1.04 (0.58-1.88,<br>p=0.890) |
| Education, years | Mean (SD) | 10.8<br>(1.1) | 0.78 (0.63-0.96,<br>p=0.017) | 0.78 (0.60-1.01,<br>p=0.059) | 0.77 (0.59-0.99,<br>p=0.045) |
| Occupational class | Non-manual | 540<br>(80.0) | - | - | - |
|  | Manual | 135<br>(20.0) | 1.30 (0.76-2.22,<br>p=0.342) | 1.06 (0.50-2.26,<br>p=0.874) | 1.05 (0.50-2.23,<br>p=0.900) |
| comorbidity_w3 | Mean (SD) | 2.3<br>(1.3) | 0.95 (0.79-1.13,<br>p=0.568) | 0.94 (0.75-1.17,<br>p=0.558) | 0.90 (0.74-1.09,<br>p=0.290) |
| Depression, HADS-D | Mean (SD) | 2.8<br>(2.3) | 1.15 (1.05-1.25,<br>p=0.002) | 1.09 (0.97-1.21,<br>p=0.141) | 1.10 (0.98-1.23,<br>p=0.120) |
| BMI, kg/m2 | Mean (SD) | 27.7<br>(4.5) | 0.98 (0.93-1.04,<br>p=0.532) | 0.95 (0.88-1.01,<br>p=0.107) | 0.95 (0.90-1.01,<br>p=0.088) |
| Physical activity level | High | 124<br>(18.5) | - | - | - |
|  | Low | 209<br>(31.1) | 1.40 (0.74-2.65,<br>p=0.295) | 1.27 (0.59-2.75,<br>p=0.539) | 1.24 (0.57-2.71,<br>p=0.580) |
|  | Moderate | 338<br>(50.4) | 0.89 (0.48-1.65,<br>p=0.705) | 0.82 (0.41-1.65,<br>p=0.582) | 0.84 (0.42-1.67,<br>p=0.610) |
| Smoking history | Never | 354<br>(51.8) | - | - | - |
|  | Current | 44<br>(6.4) | 1.80 (0.77-4.24,<br>p=0.178) | 1.45 (0.46-4.64,<br>p=0.527) | 1.39 (0.42-4.61,<br>p=0.590) |
|  | Ex-smoker | 286<br>(41.8) | 0.96 (0.60-1.52,<br>p=0.862) | 1.17 (0.66-2.04,<br>p=0.594) | 1.14 (0.66-1.96,<br>p=0.650) |

## Notes

**Funding:** The present manuscript received no direct funding. DSM is undertaking a PhD Clinical Research Fellowship funded by the Masonic Charitable Foundation and the Royal College of Psychiatrists, United Kingdom. DSM and TCR are members of the Alzheimer Scotland Dementia Research Centre, which is funded by Alzheimer Scotland. All researchers are independent of their funders. Age UK’s Disconnected Mind project supported data collection for the LBC1936 study, the UK’s Biotechnology and Biological Sciences Research Council (BBSRC) and the Economic and Social Research Council (BB/W008793/1). The Lothian Birth Cohort 1936 study acknowledges the financial support of NHS Research Scotland (NRS) through the Edinburgh Clinical Research Facility. DAG is supported by funding from the Wellcome Trust 4-year PhD in Translational Neuroscience: training the next generation of basic neuroscientists to embrace clinical research [108890/Z/15/Z].

**Conflicts of interest:** The authors have stated explicitly that there are no conflicts of interest in connection with this article. DAG is a part-time employee of Optima partners, a health data science consultancy company. Optima had no role or influence in this study.

### Competing Interest Statement

The authors have stated explicitly that there are no conflicts of interest in connection with this article. DAG is a part-time employee of Optima partners, a health data science consultancy company. Optima had no role or influence in this study.

### Funding Statement

The present manuscript received no direct funding. DSM is undertaking a PhD Clinical Research Fellowship funded by the Masonic Charitable Foundation and the Royal College of Psychiatrists, United Kingdom. DSM and TCR are members of the Alzheimer Scotland Dementia Research Centre, which is funded by Alzheimer Scotland. All researchers are independent of their funders. Age UK Disconnected Mind project supported data collection for the LBC1936 study, the UK Biotechnology and Biological Sciences Research Council (BBSRC) and the Economic and Social Research Council (BB/W008793/1). The Lothian Birth Cohort 1936 study acknowledges the financial support of NHS Research Scotland (NRS) through the Edinburgh Clinical Research Facility. DAG is supported by funding from the Wellcome Trust 4-year PhD in Translational Neuroscience: training the next generation of basic neuroscientists to embrace clinical research [108890/Z/15/Z].

### Author Declarations

All data is available from https://www.ed.ac.uk/lothian-birth-cohorts/data-access-collaboration

